# Fetal Growth Associated with Maternal Rheumatoid Arthritis and Juvenile Idiopathic Arthritis

**DOI:** 10.1101/2024.02.29.24303573

**Authors:** Eugenia Y. Chock, Zeyan Liew, Lars Henning Pedersen, Mette Østergaard Thunbo

**Affiliations:** Section of Rheumatology, Allergy and Immunology, Yale School of Medicine, New Haven, CT; Department of Environmental Health Sciences, and Yale Center for Perinatal, Pediatric, and Environmental Epidemiology, Yale School of Public Health, New Haven, USA; Department of Obstetrics and Gynecology, Clinical Medicine, Aarhus University and Aarhus University Hospital, Aarhus, Denmark; Department of Clinical Medicine, Aarhus University, Denmark

## Abstract

**Background:** Prior studies indicated that women with rheumatoid arthritis (RA) and juvenile idiopathic arthritis (JIA) are at twice higher risk of developing adverse pregnancy outcomes, this include preterm births and infants with low birth weight. A wide knowledge gap exists in our current understanding of how RA and JIA affect fetal growth during pregnancy.

**Objective:** We aimed to evaluate fetal growth among patients with RA/JIA by comparing fetal growth indicators of offspring born to this population, compared to individuals without RA/JIA. We hypothesized that fetal growth among patients with RA/JIA is reduced, compared to individuals without RA/JIA.

**Study Design:** We conducted a population-based cohort study in Denmark from 2008-2018 which included 503,491 individuals with singleton pregnancies. Among them, 2,206 were patients with RA and JIA. We linked several nationwide databases and clinical registries in Denmark to achieve our aim. Through the Danish Fetal Medicine Database (DFMD), we obtained fetal biometric measurements gathered from second trimester fetal ultrasound scans. We used International Classification of Diseases (ICD)-10 codes to identify pregnant patients with RA/JIA from the Danish National Patient Registry and linked them to the DFMD, other variables of interest were obtained from different Danish clinical registries. Next, we computed fetal growth gradient between second trimester and birth, using the mean difference in Z-score distances for each fetal growth indicator. We also estimated the risk of small for gestational age (SGA), all outcomes were compared between pregnant individuals with and without RA/JIA and adjusted for confounders.

**Results:** Maternal RA and JIA was not associated with a reduction of estimated fetal weight (EFW) at mid-pregnancy [adjusted mean EFW Z-score difference of 0.05 (95% CI 0.01, 0.10; *p*=0.022)], but lower birth weights were observed among offspring [adjusted mean Z-score difference of -0.08 (95% CI -0.13, -0.04; *p*<0.001)]. We observed reduced mean Z-score differences in weight gradient from second trimester to birth among offspring of patients with RA/JIA who used corticosteroids [-0.26 (95% CI -0.11, -0.41; *p*<0.001)], and sulfasalazine [-0.61 (95% CI -0.45, -0.77; *p*<0.001)] during pregnancy. Maternal RA/JIA was also associated with SGA [aOR of 1.47 (95% CI 1.16, 1.83; *p*<0.001)]. Similarly, the risk estimates were higher among corticosteroid [aOR 3.44 (95% CI 2.14, 5.25; *p*<0.001)] and sulfasalazine [(aOR 2.28 (95% CI 1.22, 3.88; *p*=0.005)) users.

**Conclusion:** Among pregnant patients with RA/JIA, fetal growth restriction may be most apparent after 18 to 22 weeks of gestational age. Closer antenatal monitoring around this period should be considered for this population.

## Introduction

Rheumatoid arthritis (RA) and juvenile idiopathic arthritis (JIA) are autoimmune inflammatory arthritis that disproportionately affect females.^1^ Patients with rheumatoid arthritis and juvenile idiopathic arthritis (RA/JIA) have worse pregnancy and birth outcomes compared to those without the conditions. ^2–4^ Offspring of patients with RA/JIA are at roughly double the risk of being born preterm; furthermore, infants born to patients with RA are likely to be smaller for gestational age (SGA), although this is observed at a lesser degree among patients with JIA.^2,4–7^ Offspring impacted by fetal growth restriction may go on to develop short stature and chronic medical conditions during their adult life. ^8–11^ Placental insufficiency is one of the common causes of reduced fetal growth and low birth weight.^12^ RA has been implicated to directly affect placental growth and function, ^13,14^ although other factors such as medication use and disease activity may also contribute to smaller-sized offspring.^13,15^ Prior fetal growth studies among patients with RA focused on birth weights and maternal cytokines influencing fetal growth. ^16^ Our study would be the first that details actual fetal growth measurements in utero and provide insights to fetal growth trajectories in this population. We aim to compare fetal and birth weights of offspring born to individuals with and without RA/JIA. Understanding the fetal growth pattern will help clinicians better identify vulnerable periods during pregnancy which may impact fetal growth in this population. We also investigated antirheumatic therapy (ART) uptake during pregnancy, in relation to fetal growth patterns.

## Materials and Methods

### Data Source

We conducted a population-based cohort study using several administrative and clinical registries in Denmark from years 2008 – 2018. Particularly, we linked the Danish Fetal Medicine Database (DFMD), the Danish Medical Birth Registry (DMBR), the Danish National Patient Registry (DNPR), and the Danish National Prescription Registry (NPR), through unique civil registration number assigned for each Danish resident. The linked registries allow for identification of medical and sociodemographic records of mother-offspring pairs.

### Identification of Study Population and Patient Characteristics

Our study consisted of pregnant individuals identified from DFMD, who had singleton pregnancies at 22 weeks gestational age and further (Figure 1). We used ICD-10 codes to identify patients who were diagnosed with RA/JIA 10 years prior, or during their pregnancies from the DNPR (Table S1), the positive predictive value of RA diagnosis within the DNPR is 79%.^17^ Pregnant individuals who did not have ICD-10 codes of RA and JIA served as the comparator (unexposed) group.

**Figure 1.**
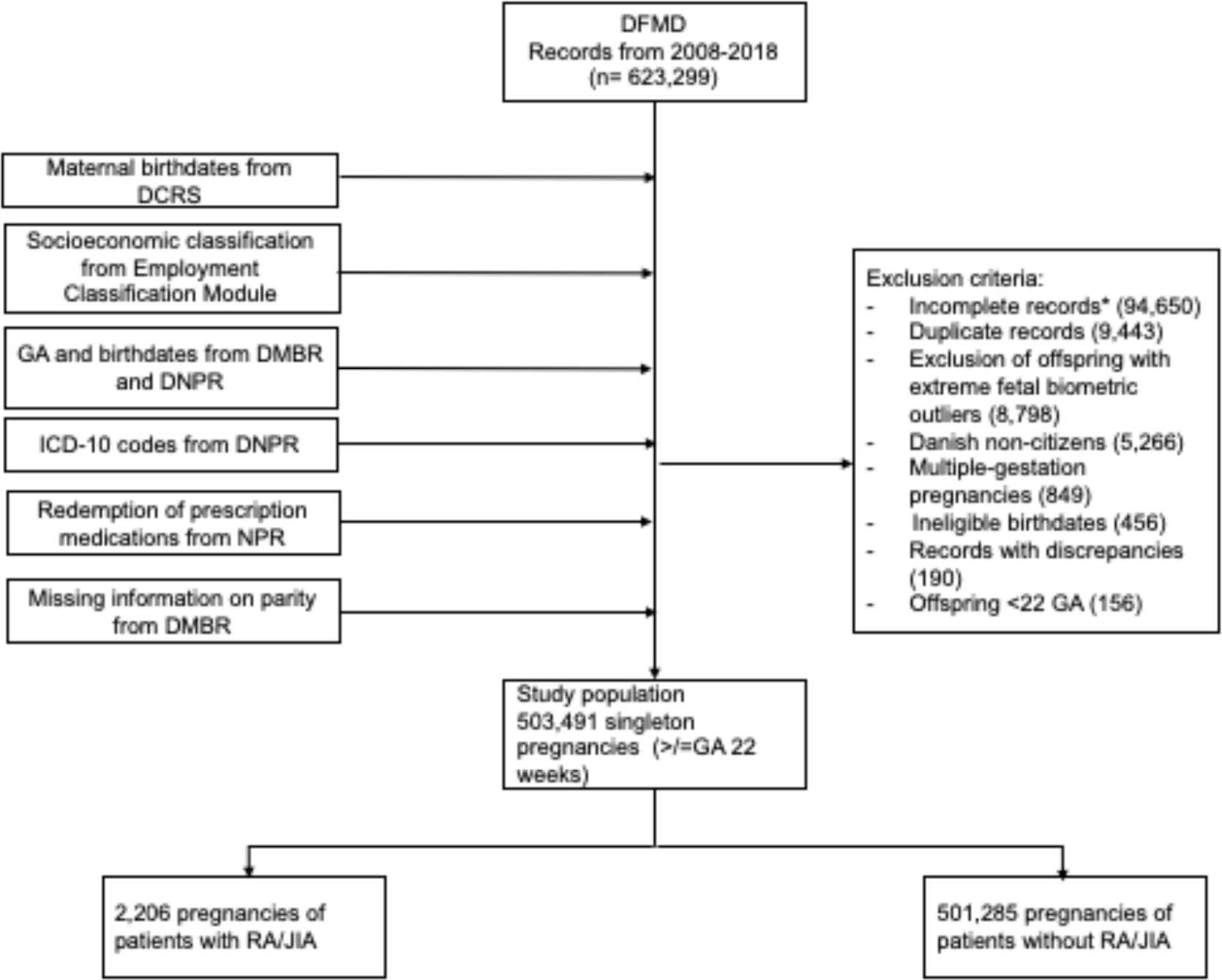
Flowchart of patient selection from linked clinical registries. **Legends**: DFMD: Danish Fetal Medicine Database; DCRS: Danish Civil Registration System; DMBR: Danish Medical Birth Registry; DNPR: Danish National Patient Registry; NPR: National Prescription Registry; RA: rheumatoid arthritis; JIA: juvenile idiopathic arthritis. * Incomplete records: pregnancy and birth records without gestational ages, lack of second trimester fetal ultrasound information, lack of birth weights.

### Antirheumatic Therapy Uptake Before and After Second Trimester Fetal Ultrasound Study

We identified the ART of interest with Anatomical Therapeutic Chemical (ATC) codes (Table S2) from the NPR. A patient with RA/JIA was considered exposed to an ART when at least one refill of ART during pregnancy was recorded, defined as 30 to 60 days’ worth of medication. We described the uptake of ART before and after second trimester fetal ultrasound studies (Figure 2).

**Figure 2.**
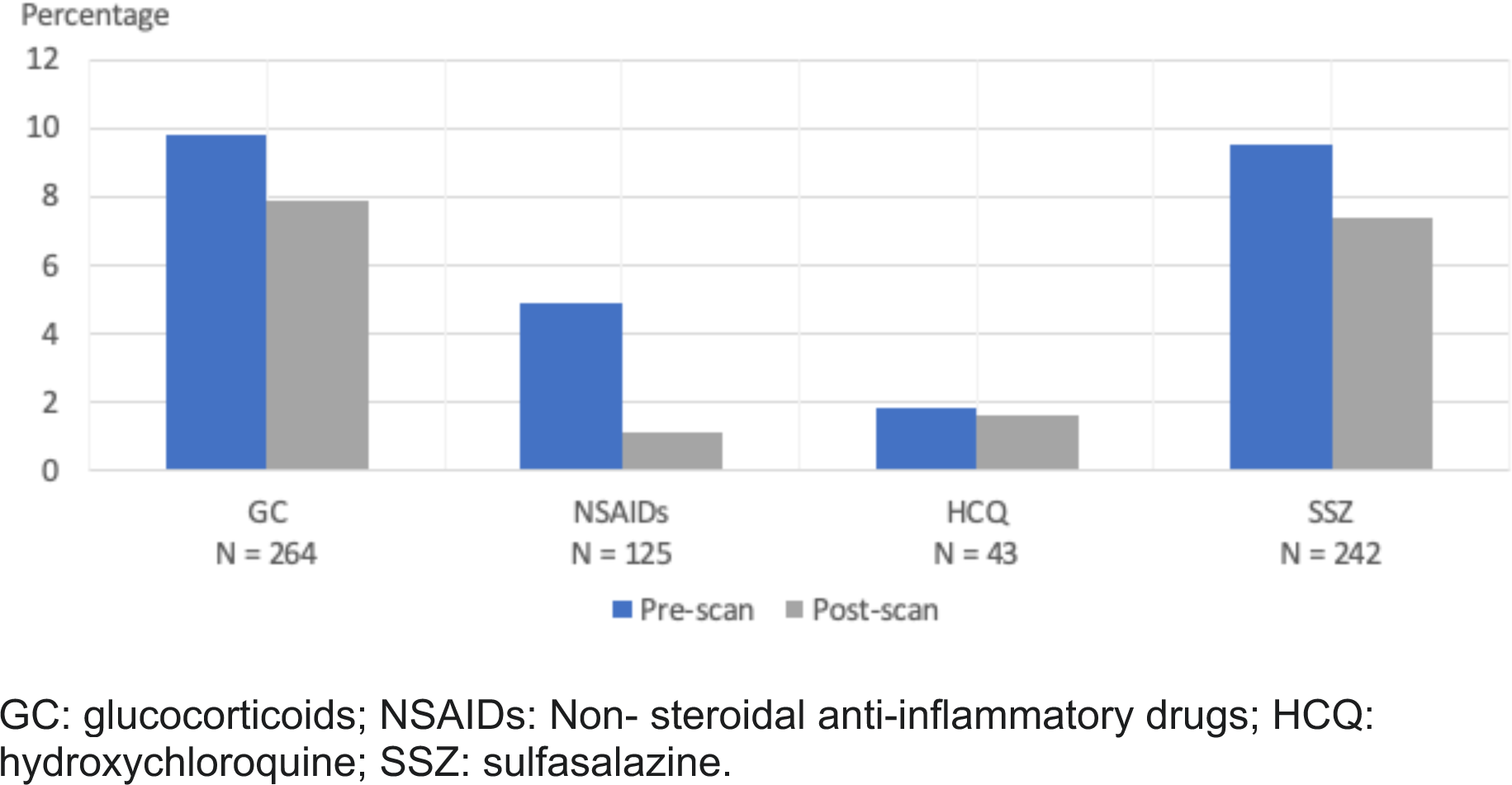
Proportion of antirheumatic therapy uptake before and after second trimester fetal ultrasound. GC: glucocorticoids; NSAIDs: Non- steroidal anti-inflammatory drugs; HCQ: hydroxychloroquine; SSZ: sulfasalazine.

### Ascertainment of Outcomes

#### Fetal and Birth Weights

The DFMD contains standard fetal biometric measurements obtained during first and second trimester fetal ultrasound examinations of nearly all pregnant individuals in Denmark.^18^ First and second trimester fetal ultrasounds are expected to have >90% completeness covering all pregnant individuals in Denmark since 2008.^18^ In Denmark, all fetal ultrasounds are performed by trained and certified sonographers following standard operating procedures, free of charge. We extracted fetal head circumferences, abdominal circumferences, and femur lengths (all in mm) from second trimester (gestational weeks 18-22) fetal ultrasounds of the study population. We then computed the estimated fetal weights (EFW) with the Hadlock formula based on those measurements.^19^ We anticipated mostly accurate EFW estimations which are within 10% error based on prior studies. ^20,21^ We subsequently obtained birth weights of respective offspring following the second trimester fetal ultrasound.

#### Head circumferences (HC)

We extracted measurements of offspring HC from second trimester and birth to assess the direction of head growth in relation to weight.

#### Small-for-gestational-age (SGA)

Defined as birth weight at least 2 standard deviations below the mean for gestational age (corresponding to -2 Z-scores), ^22^ as an indicator of restricted fetal growth.

For all fetal and birth outcomes, we first compared pregnancies with and without RA/JIA. Next, we stratified the RA/JIA group by different Antirheumatic Therapies (ART) used during pregnancy.

### Covariates

Relevant covariates associated with outcomes of interest were selected a priori, via construction of the Direct Acyclic Graph and literature review. All analyses for outcomes were adjusted for maternal age at conception, pre-pregnancy BMI, smoking status, income status, race, parity, birth year, pre-pregnancy hypertension, pre-pregnancy diabetes, and co-medication use. Sources of several covariates were derived from registries noted in Fig. 1. As Denmark has an extensive social security system and high-income equality, we consider income status obtained from the Employment Classification Module (ECM) as an indicator of socioeconomic status. The ECM records main employment activities of Danish residents within each calendar year. ^23^ Co-medications are inclusion of dispensed medications not listed in Table S2; in addition, hospital-administered medications were excluded as the records are not available within the NPR. All other variables such as smoking status were derived from the DFMD.

### Statistical Analysis

We performed descriptive statistics to report general characteristics of the study population. We then converted EFW and HC to Z-scores using means and standard deviations from international and published standardized growth measures stratified by gestational week.^19,24^ To understand the direction of fetal growth, we calculated the gradient of weight and HC differences between second trimester and birth of the offspring. The formula to calculate gradient in weight and HC differences between second trimester and at birth was as follows:

Negative values indicate reduced growth between second trimester and birth, compared to offspring born to individuals without RA/JIA. We performed multivariable linear regression analyses to estimate the mean differences in fetal weight, birth weight, weight gradient, and head circumference Z scores, according to maternal RA/JIA diagnoses and their ART use during pregnancy. We conducted multiple logistic regression to estimate the risk ratio and 95% confident interval for SGA among offspring associated with maternal RA/JIA and the respective maternal ART exposures. All analyses were adjusted for confounding variables.

### Mediation Analysis

Finally, we performed causal mediation analysis to evaluate the degree to which pre- eclampsia (PEC) mediates the association between pregnancy exposed to RA/JIA and risk of SGA among offspring, adjusted for covariates noted above. We used the *CMAverse* package in RStudio to perform mediation analysis. In the mediation analysis model, we estimated the Odds Ratio (OR) of total effect and controlled direct effect between exposure and outcome.

All analyses were performed with RStudio version 2022.12.0.

## Results

Our study population included 503,491 individuals with singleton pregnancies (study selection process depicted in Figure 1). In total, there were 2,206 patients with RA/JIA, comprising of 86.0% RA (N= 1,897) and 26.7% (N= 588) JIA patients. Compared to pregnant individuals without RA/JIA, patients with the conditions were more likely to be older than 35 years of age at the time of pregnancy (21.7% vs. 16.8%), had BMI between 25 to <30 (24.2% vs. 21.8%), and experienced more pre-term births born during 32 and <37 weeks gestational age (5.8% vs 4.1%). Patients with RA/JIA were likelier to have co-morbidities before and during pregnancy, this includes pre-gestational hypertension (1.9% vs 0.9%), pre-gestational diabetes (1.2% vs 0.7%), gestational hypertension (3.1% vs 2.2%), pre-eclampsia and eclampsia (4.9% vs 3.9%). 25.5% of patients with RA/JIA underwent C-section compared to those without (20.0%). Patients with RA/JIA were also of lower socioeconomic status (28.9% on transfer payment and in early retirement vs 22.7%). The proportion of women with gestational diabetes were the same for both groups (3.6%). Both groups were also similar in terms of race (> 90% Caucasian) and smoking status (non-smoker 89.8% vs 89.4%) (Table 1).

**Table 1.** Baseline characteristics of study population.

| Characteristics* | Overall<br>(N=503,491) | No RA and JIA<br>(N=501,285) | RA and JIA<br>(N=2,206) |
| --- | --- | --- | --- |
| **Age (years)** | N (%) | N (%) | N (%) |
| <25 | 76,972 (15.3) | 76,608 (15.3) | 364 (16.5) |
| 25-29 | 175,596 (34.9) | 174,943 (34.9) | 653 (29.6) |
| 30-34 | 166,247 (33.0) | 165,536 (33.0) | 711 (32.2) |
| ≥35 | 84,676 (16.8) | 84,198 (16.8) | 478 (21.7) |
| **Race** |  |  |  |
| Afro-Caribbean | 5,785 (1.2) | 5,770 (1.2) | 15 (0.7) |
| Asian | 17,230 (3.5) | 17,189 (3.5) | 41 (1.9) |
| Caucasian | 457,274 (93.6) | 455,221 (93.6) | 2,053 (96.1) |
| Other <sup>±</sup> | 8,436 (1.7) | 8,409 (1.7) | 27 (1.3) |
| **BMI (kg/m<sup>2</sup>)** |  |  |  |
| <18.5 | 31,927 (6.5) | 31,798 (6.5) | 129 (6.1) |
| 18.5-24 | 287,290 (58.8) | 286,083 (58.8) | 1,207 (56.7) |
| 25-29 | 106,537 (21.8) | 105,022 (21.8) | 515 (24.2) |
| ≥30 | 62,884 (12.9) | 62,60 (12.9) | 277 (13.0) |
| **Smoking status during pregnancy** |  |  |  |
| Former | 7,642 (1.5) | 7,606 (1.5) | 36 (1.7) |
| No | 442,531 (89.4) | 440,591 (89.4) | 1,940 (89.8) |
| Yes | 44,832 (9.1) | 44,648 (9.0) | 184 (8.5) |
| **Employment status** |  |  |  |
| In school | 33,471 (6.7) | 33,317 (6.6) | 154 (7.0) |
| Employed | 351,448 (69.8) | 350,043 (69.8) | 1,405 (63.7) |
| Transfer payment¶ | 111,390 (22.1) | 110,811 (22.1) | 579 (26.2) |
| Early retirement | 3,251 (0.6) | 3,191 (0.6) | 60 (2.7) |
| Unemployed | 3,699 (0.7) | 3,691 (0.7) | 8 (0.4) |
| **Parity** |  |  |  |
| Nulliparous | 180,161 (35.8) | 179,317 (35.8) | 844 (38.3) |
| Multiparous | 323,324 (64.2) | 321,962 (64.2) | 1,362 (61.7) |
| **Co-morbidities** |  |  |  |
| Pregestational hypertension | 4,472 (0.9) | 4,431 (0.9) | 41 (1.9) |
| Gestational hypertension | 10,856 (2.2) | 10,778 (2.2) | 68 (3.1) |
| Pre-eclampsia/eclampsia | 19,814 (3.9) | 19,707 (3.9) | 107 (4.9) |
| Pregestational diabetes | 3,611 (0.7) | 3,585 (0.7) | 26 (1.2) |
| Gestational diabetes | 18,031 (3.6) | 17,951 (3.6) | 80 (3.6) |
| **Birth- related characteristics** |  |  |  |
| **Pregnancy Outcome** |  |  |  |
| Stillborn | 1,049 (0.2%) | 1,045 (0.2%) | 4 (0.2%) |
| **Gestational Age at Birth (weeks)** |  |  |  |
| <32 | 3,606 (0.7%) | 3,587 (0.7%) | 19 (0.8%) |
| =/>32 to <37 | 20,488 (4.1%) | 20,361 (4.1%) | 127 (5.8%) |
| =/>37 to <42 | 464,218 (92.2%) | 462,218 (92.2%) | 2,000 (90.7%) |
| =/> 42 | 15,179 (3.0%) | 15,119 (3.0%) | 60 (2.7%) |
| **Mode of Delivery** |  |  |  |
| Vaginal delivery | 402,700 (80.0%) | 401,056 (80.0%) | 1,644 (74.5%) |
| C-section | 100,791 (20.0%) | 100,229 (20.0%) | 562 (25.5%) |

\* Variables contain missing values.
¶ Transfer payment: Temporary income for individuals administered by employer or government due to a health condition. <sup>42</sup>
± Identify as multiracial

When assessing the EFW Z-scores obtained from fetal US biometric measurements during 2nd trimester (Table S2), the overall adjusted mean EFW Z-score difference among offspring of patients with RA/JIA was 0.05 compared to those unexposed (95% CI 0.01, 0.10; *p*=0.022) (Table S3). When stratified by ART, patients with RA/JIA who used sulfasalazine (SSZ) had an adjusted mean EFW Z-score difference of 0.38 (95% CI 0.24, 0.50; *p*<0.001). For birth weight, the overall adjusted mean Z-score difference among offspring of patients with RA/JIA was -0.08 (95% CI -0.13, -0.04; *p*<0.001) (Table S3). The adjusted mean Z-score difference for women with RA/JIA who used corticosteroids (CS) during pregnancy was -0.31 (95% CI -0.43, -0.18; *p*<0.001); and -0.23 (95% CI -0.37, -0.10; *p*<0.001) among SSZ users, compared to those unexposed (Table S3).

When assessing fetal growth by analyzing the gradient in mean differences of weight and head circumferences between 2nd trimester and birth, the adjusted Z-score mean difference between EFW and birth weight (from 2nd trimester US) was -0.14 (95% CI - 0.08, -0.19; *p*<0.001) for offspring of patients with RA/JIA compared with no RA/JIA (Figure 3). Patients with RA/JIA who used CS during pregnancy had an adjusted Z-score EFW and birth weight difference of -0.26 (95% CI -0.11, -0.41; p<0.001), -0.25 (95%CI -0.64, 0.13; *p*=0.2) for hydroxychloroquine (HCQ) users, and -0.61 (95%CI - 0.45, -0.77; *p*<0.001 for SSZ use during pregnancy, compared with no RA/JIA (Figure 3). The overall adjusted Z-score difference of head circumferences between 2nd trimester and birth was -0.03 (95%CI -0.09, 0.02; *p*=0.2) comparing RA/JIA to no RA/JIA. The adjusted Z-score differences in head circumferences between 2nd trimester and birth for mothers with RA/JIA and who took CS during pregnancy was - 0.19 (95% CI -0.02, -0.36; *p*=0.026), and -0.33 (95% CI -0.74, 0.09; *p*=0.12) for HCQ, and -0.20 (95%CI -0.02, -0.37; *p*=0.03) for SSZ users during pregnancy (Figure 4).

**Figure 3.**
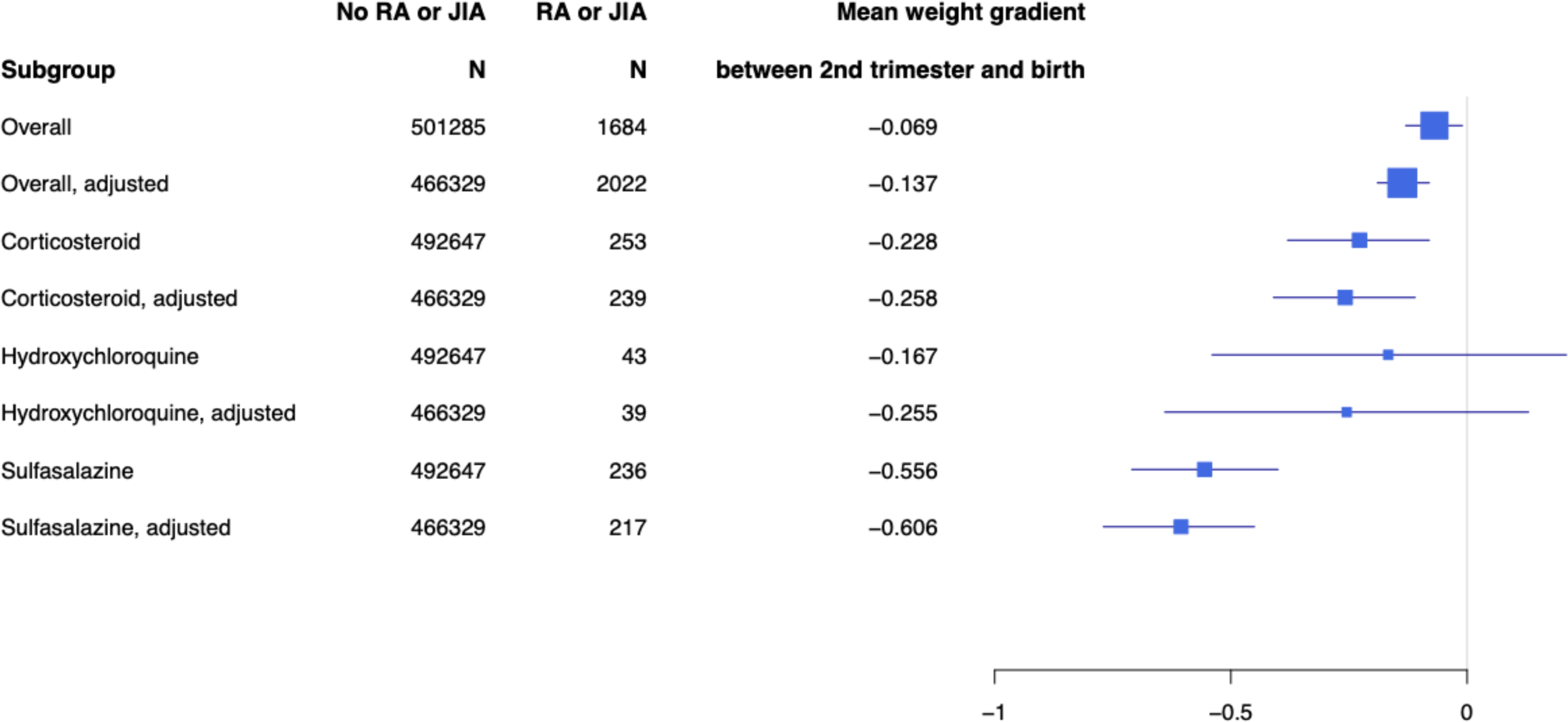
Weight gradient of offspring born to patients with RA and JIA between second trimester and at birth.

**Figure 4.**
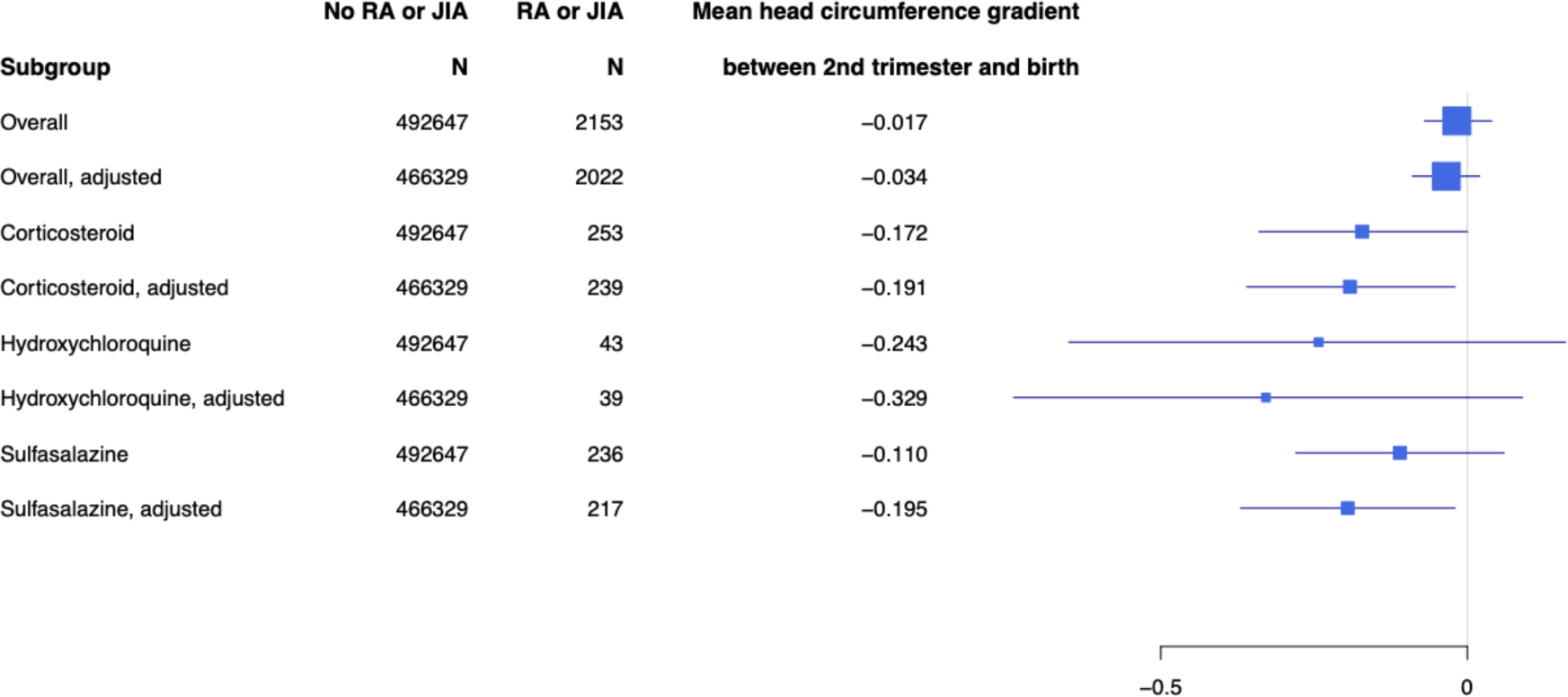
Head circumference gradient of offspring born to patients with RA and JIA between second trimester and at birth.

Offspring born to patients with RA/JIA were associated with a higher risk of SGA compared with no RA/JIA (aOR 1.47, 95%CI 1.16, 1.83; *p*<0.001) (Table 2). The adjusted OR for SGA stratified by ART use among patients with RA/JIA were as follows: CS [aOR 3.44 (95% CI 2.14,5.25; *p*<0.001)], HCQ [aOR 3.12 (95%CI 0.74, 8.80; *p*=0.062)], SSZ [aOR 2.28 (95%CI 1.22, 3.88; *p*=0.005)]. In mediation analysis, preeclampsia was estimated to mediate about 4.3% of the total effect between prenatal RA/JIA exposure and offspring risk for SGA (Table S4).

**Table 2.** Risk estimates of small for gestational age infants among patients with RA and JIA, stratified by antirheumatic therapy use during pregnancy.

| Patient subgroups | Proportion with SGA (%) | Crude model |  |  | Adjusted model |  |  |
| --- | --- | --- | --- | --- | --- | --- | --- |
|  |  | Odds Ratio (OR) | 95% CI | *p*-value | OR | 95% CI | *p*-value |
| No RA and JIA | 13,025/<br>501,285 (2.6) | 1.00 | Reference |  | 1.00 | Reference |  |
| RA and JIA | 87 / 2,206 (3.9) | 1.54 | 1.23, 1.90 | **<0.001** | 1.47 | 1.16, 1.83 | **<0.001** |
| - CS | 24 / 264 (9.1) | 3.75 | 2.40, 5.58 | **<0.001** | 3.44 | 2.14, 5.25 | **<0.001** |
| - HCQ | 3 /43 (7.0) | 2.81 | 0.68, 7.74 | 0.084 | 3.12 | 0.74, 8.80 | 0.062 |
| - SSZ | 14 / 242 (5.8) | 2.30 | 1.28, 3.80 | **0.002** | 2.28 | 1.22, 3.88 | **0.005** |

\* Adjusted for maternal age, body mass index, smoking, income status, race, parity, and birth year. CS: Corticosteroids; HCQ: hydroxychloroquine; SSZ: sulfasalazine

## Structured Discussion

### Principal Findings

To our knowledge, this is the first population-based cohort study which investigated fetal growth with fetal ultrasound biometric data among offspring of patients with RA and JIA. We found that EFW among offspring of patients with RA/ JIA during the second trimester were slightly higher compared to those without RA/JIA, notably EFW were highest among SSZ users during pregnancy. Interestingly, the finding did not persist up to birth as birth weights among offspring of patients with RA/JIA were lower than the non-RA/JIA group. We also found simultaneous reduction in head growth in these offspring on analysis of head circumferences between second trimester and birth. The greatest reduction in fetal growth gradient between second trimester and birth were among RA/JIA patients who used CS and SSZ during pregnancy. It is likely that RA/JIA patients who were taking CS and SSZ during pregnancy had more active clinical disease activity, necessitating their use. We observed a modest drop in the uptake of both medications after second trimester fetal ultrasounds were performed (Figure 2), indicating that the reduction in fetal growth may be exposure dependent and cessation of use is unlikely to reverse the phenomenon.

Finally, we also found that patients with RA/JIA were likelier to deliver infants who are SGA, compared to those without RA/JIA. Consistent with our findings on fetal weight and head circumference reduction between second trimester and birth, infants of patients with RA/JIA who used CS and SSZ were at highest risk of being born SGA. Previous studies have demonstrated the negative impact of CS exposure on fetal biometric measurements. ^25^ Although higher proportion of RA/JIA patients experience PEC, PEC only weakly mediated the total effect of RA/JIA on SGA.

## Results

Our results are consistent with prior studies on pregnancy outcomes among patients with RA/JIA: pregnant patients with RA tend to be older at the time of pregnancy, at higher risk for hypertensive disorders during pregnancy, tend to undergo C-section deliveries, and their offspring are likelier to have restricted fetal growth. ^2,15,26^

Interestingly, our study found that patients with RA/JIA who used CS or SSZ during pregnancy had higher EFW during the second trimester compared to fetuses of patients without RA/JIA (Table S3). As patients with RA/JIA are at higher risk of early pregnancy losses and subfertility, ^27,28^ it is plausible that the finding of higher EFW during second trimester among women with RA/JIA was influenced by survivor bias. This is a situation in which offspring exposed to maternal RA/JIA were not likely to survive until birth and many pregnancy losses may have already occurred during early pregnancy. ^29^ Despite the overall higher EFW among patients with RA/JIA in our study, the pattern did not persist up to birth as infants born to these groups also experienced slower growth after second trimester, compared to infants born to patients without RA/JIA. The findings on CS use during pregnancy among patients with RA/JIA were concurrent with studies done on the general population, in that antenatal CS use is highly associated with reduced fetal growth and low birth weight. ^30,31^ The gradient of fetal growth from second trimester to birth was most reduced among RA/JIA patients who used SSZ during pregnancy. Of note, SSZ is generally recommended for the treatment of RA during pregnancy, it had shown promise in the treatment of PEC and pre-term labor in both *in vivo* and *in vitro* studies. ^13,32^ Therefore this finding of reduced birth weight among SSZ users remains unclear. A plausible explanation would be that high RA/JIA clinical disease activity, which had necessitated the use of CS and SSZ, is likely responsible for the negative effect on fetal growth. This is supported by prior studies which reported an association between adverse pregnancy outcomes and high clinical disease activity in patients with inflammatory arthritis. ^7,15,33,34^

Our study also had several findings that have been similarly reported in other studies on this population: patients with RA/JIA in our study have higher rates of C-sections and pre-eclampsia; ^6,26,28,35^ however the rates of stillborn are the same as individuals without RA/JIA. ^6^ From our mediation analysis, we found that PEC accounted for only about 4% of fetal growth restriction.

## Clinical Implications

We postulate that the period of pregnancy after 18-22 gestational age may be most vulnerable for optimal growth and development among offspring of pregnant RA/JIA patients. Therefore, antenatal factors during this critical period would be important to examine. Although PEC may have a minor mediating role in SGA within this population; other major factors include antenatal use of CS and clinical disease activity of RA/JIA during this period. RA/JIA patients who experience high clinical disease activity or flares are usually treated with CS. ^36,37^ As our study findings are concurrent with prior investigations on the negative impact of antenatal CS use against fetal growth, treating physicians should make every effort to control clinical disease activity during early pregnancy and minimize antenatal CS use. It is also possible that among pregnant patients with RA/JIA, cytokine interactions are occurring at the feto-maternal interface, resulting in placental dysfunction and leading to decreased fetal weight during later part of pregnancy.^13,38^ We did not find a meaningful association between antenatal HCQ use with the outcomes of interest, but the number of RA/JIA on HCQ during pregnancy were small (N=43).

Closer monitoring and follow-ups between second trimester and birth may be considered in routine antenatal care of patients with RA/JIA, this may include third trimester fetal ultrasound studies and monitoring of RA/JIA disease activity. As this is an observational study, further research strategies using randomized controlled trial or target trial emulation would illuminate the effect of medication use vs disease activity on fetal growth in pregnancies of patients with inflammatory arthritis. In addition, an interdisciplinary approach with clear communication between the treating rheumatologist and obstetrician is encouraged. As suggested by prior studies, our findings further solidify the recommendation that treating rheumatologists should confer with their RA/JIA patients with regards to pregnancy planning and management of disease activity during early pregnancy. ^36,39^

## Research Implications

Our study provided insight on fetal growth at two pregnancy time points, namely the second trimester and at birth. Further fetal biometric measurements via ultrasound during first and third trimester of pregnancy would provide more accurate fetal growth trajectories among patients with RA/JIA. This is critical to identify the time point at which fetal growth is most compromised during pregnancy, and further aid in investigating maternal or placental factors that occurred during that specific period. We also need granular information around when fetal ultrasound is performed, such as RA/JIA disease activity and biologic ART use.

Our findings also open the door for further mechanistic studies on the role of placental growth and development among patients with RA/JIA. As majority of fetal weight gain occurs during the third trimester of pregnancy, both internal and external insults among patients with RA/JIA may result in placental insufficiency at the later stages of pregnancy, hence compromising fetal development during that period. ^40,41^

## Strengths and Limitations

This is a large, population-based cohort study which included all singleton pregnancies ≥ 22 weeks gestational age born in Denmark during the 11-year period from 2008 to 2018. A prior study by Rom et. al. reported growth measurements at birth among offspring born to parents with RA, ^16^ our study provided more details on fetal growth measurements obtained via fetal ultrasound studies during the second trimester. We also examined ART use before and after second trimester fetal ultrasound studies and investigated if fetal growth differed by ART use, we used medication refill records instead of prescription records. Furthermore, we performed mediation analysis to understand the excess risk of SGA among RA/JIA patients that may be contributed by PEC.

Our study has several limitations, this include incomplete ascertainment of ART used during pregnancy among patients with RA/JIA, the NPR did not contain information on biologic disease modifying antirheumatic drugs prescribed to patients. We were also unable to correlate the use of CS with clinical disease activity measurements of these patients, therefore we are uncertain if antenatal CS use were indicated for RA/JIA flares or other reasons. For purpose of this study, the mean differences in Z score for fetal weight and head circumferences between second trimester and birth was an optimal measurement to understand growth direction between two time points. As we include additional fetal ultrasound measurements in future studies, a hierarchical Bayesian model may be best suited to investigate the effect of maternal exposures on fetal growth, while taking into account repeated measurements of the same individual fetus.

## Conclusion

Our study demonstrated that fetal growth restriction may be most apparent after 18 to 22 weeks of gestational age among pregnant patients with RA/JIA. A reduction in fetal growth (head circumferences and birth weights) ensued post second trimester ultrasound, resulting in smaller infants among patients with RA/JIA. This suggests that the second half of pregnancy may be a vulnerable period for optimal fetal growth among pregnant patients with RA/JIA; or that placental changes affect fetal growth in the later part of pregnancy after a certain size is achieved.

## Supporting information

Manusript

## Data Availability

Individual-level data cannot be shared publicly due to the requirements of the registry holders and the general data protection regulation in order to protect the privacy of individuals.

## References

1. Martini A, Lovell DJ, Albani S, et al. Juvenile idiopathic arthritis. Nat Rev Dis Primers. 2022;8(1):5–8. doi: 10.1038/s41572-021-00332-8.

2. Chakravarty EF, Nelson L, Krishnan E. Obstetric hospitalizations in the united states for women with systemic lupus erythematosus and rheumatoid arthritis. Arthritis Rheum. 2006;54(3):899–907. doi: 10.1002/art.21663.

3. de Man YA, Hazes JMW, van der Heide H, et al. Association of higher rheumatoid arthritis disease activity during pregnancy with lower birth weight: Results of a national prospective study. Arthritis & Rheumatism. 2009;60(11):3196–3206. 10.1002/art.24914. doi: 10.1002/art.24914.

4. Aljary H, Czuzoj-Shulman N, Spence AR, Abenhaim HA. Pregnancy outcomes in women with rheumatoid arthritis: A retrospective population-based cohort study. The Journal of Maternal-Fetal & Neonatal Medicine. 2020;33(4):618–624. 10.1080/14767058.2018.1498835. doi: 10.1080/14767058.2018.1498835.

5. Ehrmann Feldman D, Vinet É, Bernatsky S, et al. Birth outcomes in women with a history of juvenile idiopathic arthritis. J Rheumatol. 2016;43(4):804–809. doi: 10.3899/jrheum.150592.

6. Nørgaard M, Larsson H, Pedersen L, et al. Rheumatoid arthritis and birth outcomes: A danish and swedish nationwide prevalence study. J Intern Med. 2010;268(4):329–337. doi: 10.1111/j.1365-2796.2010.02239.x [doi].

7. Remaeus K, Johansson K, Askling J, Stephansson O. Juvenile onset arthritis and pregnancy outcome: A population-based cohort study. Ann Rheum Dis. 2017;76(11):1809–1814. doi: 10.1136/annrheumdis-2016-210879 [doi].

8. Barker DJ, Osmond C, Golding J, Kuh D, Wadsworth ME. Growth in utero, blood pressure in childhood and adult life, and mortality from cardiovascular disease. BMJ. 1989;298(6673):564-567. doi: 10.1136/bmj.298.6673.564.

9. Barker DJ, Bull AR, Osmond C, Simmonds SJ. Fetal and placental size and risk of hypertension in adult life. BMJ. 1990;301(6746):259-262. doi: 10.1136/bmj.301.6746.259.

10. Wang KCW, James AL, Noble PB. Fetal growth restriction and asthma: Is the damage done? Physiology (Bethesda). 2021;36(4):256–266. doi: 10.1152/physiol.00042.2020.

11. Bo S, Cavallo-Perin P, Scaglione L, Ciccone G, Pagano G. Low birthweight and metabolic abnormalities in twins with increased susceptibility to type 2 diabetes mellitus. Diabet Med. 2000;17(5):365–370. doi: 10.1046/j.1464-5491.2000.00288.x.

12. Pinney SE. Chapter 14 - metabolic disorders and developmental origins of health and disease. In: Rosenfeld CS, ed. The epigenome and developmental origins of health and disease. Boston: Academic Press; 2016:267-289. https://www.sciencedirect.com/science/article/pii/B9780128013830000141. 10.1016/B978-0-12-801383-0.00014-1.

13. Neuman RI, Smeele HTW, Jan Danser AH, Dolhain RJEM, Visser W. The sFlt-1 to PlGF ratio in pregnant women with rheumatoid arthritis: Impact of disease activity and sulfasalazine use. Rheumatology (Oxford). 2021;61(2):628–635. 10.1093/rheumatology/keab372. Accessed 11/8/2023. doi: 10.1093/rheumatology/keab372.

14. F. Förger, M. Baumann, L. Risch, et al. FRI0127 Angiogenic placental factors during pregnancy in rheumatoid arthritis. Ann Rheum Dis. 2016;75(Suppl 2):474. http://ard.bmj.com/content/75/Suppl_2/474.1.abstract. doi: 10.1136/annrheumdis-2016-eular.3520.

15. Hellgren K, Secher AE, Glintborg B, et al. Pregnancy outcomes in relation to disease activity and anti-rheumatic treatment strategies in women with rheumatoid arthritis. Rheumatology (Oxford). 2021. doi: keab894 [pii].

16. Rom AL, Wu CS, Olsen J, et al. Fetal growth and preterm birth in children exposed to maternal or paternal rheumatoid arthritis: A nationwide cohort study. Arthritis Rheumatol. 2014;66(12):3265–3273. doi: 10.1002/art.38874 [doi].

17. Ibfelt EH, Sørensen J, Jensen DV, et al. Validity and completeness of rheumatoid arthritis diagnoses in the nationwide DANBIO clinical register and the danish national patient registry. Clin Epidemiol. 2017;9:627–632. doi: 10.2147/CLEP.S141438 [doi].

18. Ekelund CK, Kopp TI, Tabor A, Petersen OB. The danish fetal medicine database. Clin Epidemiol. 2016;8:479–483. doi: clep-8-479 [pii].

19. Hadlock FP, Harrist RB, Sharman RS, Deter RL, Park SK. Estimation of fetal weight with the use of head, body, and femur measurements—A prospective study. Obstet Gynecol. 1985;151(3):333–337. https://www.sciencedirect.com/science/article/pii/0002937885902984. doi: 10.1016/0002-9378(85)90298-4.

20. Dittkrist L, Vetterlein J, Henrich W, et al. Percent error of ultrasound examination to estimate fetal weight at term in different categories of birth weight with focus on maternal diabetes and obesity. BMC Pregnancy and Childbirth. 2022;22(1):241. 10.1186/s12884-022-04519-z. doi: 10.1186/s12884-022-04519-z.

21. Benson-Cooper S, Tarr GP, Kelly J, Bergin CJ. Accuracy of ultrasound in estimating fetal weight in new zealand. Australasian Journal of Ultrasound in Medicine. 2021;24(1):13–19. 10.1002/ajum.12239. doi: 10.1002/ajum.12239.

22. Lee PA, Chernausek SD, Hokken-Koelega ACS, Czernichow P, International Small for Gestational Age Advisory Board. International small for gestational age advisory board consensus development conference statement: Management of short children born small for gestational age, april 24-october 1, 2001. *Pediatrics*. 2003;111(6 Pt 1):1253-1261. doi: 10.1542/peds.111.6.1253.

23. Svane-Petersen AC, Framke E, Sørensen JK, Rugulies R, Madsen IEH. Cohort profile: The danish work life course cohort study (DaWCo). BMJ Open. 2019;9(11):e029658–029658. doi: 10.1136/bmjopen-2019-029658.

24. Marsál K, Persson PH, Larsen T, Lilja H, Selbing A, Sultan B. Intrauterine growth curves based on ultrasonically estimated foetal weights. Acta Paediatr. 1996;85(7):843–848. doi: 10.1111/j.1651-2227.1996.tb14164.x.

25. Norberg H, Stålnacke J, Diaz Heijtz R, et al. Antenatal corticosteroids for preterm birth: Dose-dependent reduction in birthweight, length and head circumference. Acta Paediatr. 2011;100(3):364–369. doi: 10.1111/j.1651-2227.2010.02074.x.

26. Sim BL, Daniel RS, Hong SS, et al. Pregnancy outcomes in women with rheumatoid arthritis: A systematic review and meta-analysis. JCR: Journal of Clinical Rheumatology. 2023;29(1). https://journals.lww.com/jclinrheum/fulltext/2023/01000/pregnancy_outcomes_in_women_with_rheumatoid.6.aspx.

27. Marianne Wallenius, Kjell Å. Salvesen, Anne K. Daltveit, Johan F. Skomsvoll. Miscarriage and stillbirth in women with rheumatoid arthritis. J Rheumatol. 2015;42(9):1570–1572. http://www.jrheum.org/content/42/9/1570.abstract. doi: 10.3899/jrheum.141553.

28. Smeele HTW, Dolhain RJEM. Current perspectives on fertility, pregnancy and childbirth in patients with rheumatoid arthritis. Semin Arthritis Rheum. 2019;49(3, Supplement):S32-S35. https://www.sciencedirect.com/science/article/pii/S0049017219306456. doi: 10.1016/j.semarthrit.2019.09.010.

29. Liew Z, Olsen J, Cui X, Ritz B, Arah OA. Bias from conditioning on live birth in pregnancy cohorts: An illustration based on neurodevelopment in children after prenatal exposure to organic pollutants. Int J Epidemiol. 2015;44(1):345–354. doi: 10.1093/ije/dyu249.

30. Thorp JA, Jones PG, Knox E, Clark RH. Does antenatal corticosteroid therapy affect birth weight and head circumference? Obstetrics & Gynecology. 2002;99(1):101–108. https://www.sciencedirect.com/science/article/pii/S0029784401016568. doi: 10.1016/S0029-7844(01)01656-8.

31. Murphy KE, Willan AR, Hannah ME, et al. Effect of antenatal corticosteroids on fetal growth and gestational age at birth. Obstet Gynecol. 2012;119(5):917–923. doi: 10.1097/AOG.0b013e31825189dc.

32. Sykes L. Sulfasalazine augments a pro-inflammatory response in interleukin-1β- stimulated amniocytes and myocytes. Immunology. 12;146(4):630–644.

33. Remaeus K, Johansson K, Granath F, Stephansson O, Hellgren K. Pregnancy outcomes in women with psoriatic arthritis in relation to presence and timing of antirheumatic treatment. Arthritis Rheumatol. 2022;74(3):486–495. doi: 10.1002/art.41985 [doi].

34. Gerosa M, Cecilia BC, Pregnolato F, et al. Pregnancy in juvenile idiopathic arthritis: Maternal and foetal outcome, and impact on disease activity. Therapeutic Advances in Musculoskeletal. 2022;14:1759720X221080375. 10.1177/1759720X221080375. doi: 10.1177/1759720X221080375.

35. Bharti B, Lee SJ, Lindsay SP, et al. Disease severity and pregnancy outcomes in women with rheumatoid arthritis: Results from the organization of teratology information specialists autoimmune diseases in pregnancy project. J Rheumatol. 2015;42(8):1376–1382. http://www.jrheum.org/content/42/8/1376.abstract. doi: 10.3899/jrheum.140583.

36. Sammaritano LR, Bermas BL, Chakravarty EE, et al. 2020 american college of rheumatology guideline for the management of reproductive health in rheumatic and musculoskeletal diseases. Arthritis Care Res (Hoboken). 2020;72(4):461-488. doi: 10.1002/acr.24130.

37. Fraenkel L, Bathon JM, England BR, et al. 2021 american college of rheumatology guideline for the treatment of rheumatoid arthritis. Arthritis Rheumatol. 2021;73(7):1108- 1123. doi: 10.1002/art.41752 [doi].

38. Atta DS, Girbash EF, Abdelwahab SM, Abdeldayem HM, Tharwat I, Ghonaim R. Maternal cytokines and disease severity influence pregnancy outcomes in women with rheumatoid arthritis. The Journal of Maternal-Fetal & Neonatal Medicine. 2016;29(20):3358–3363. 10.3109/14767058.2015.1127342. doi: 10.3109/14767058.2015.1127342.

39. CastroGutierrez A, Young K, Bermas BL. Pregnancy and management in women with rheumatoid arthritis, systemic lupus erythematosus, and obstetric antiphospholipid syndrome. Med Clin North Am. 2021;105(2):341–353. https://ovidsp.ovid.com/ovidweb.cgi?T=JS&CSC=Y&NEWS=N&PAGE=fulltext&D=emexa&AN=2011008387. http://wa4py6yj8t.search.serialssolutions.com/?url_ver=Z39.88-2004&rft_val_fmt=info:ofi/fmt:kev:mtx:journal&rfr_id=info:sid/Ovid:emexa&rft.genre=article&rft_id=info:doi/10.1016%2Fj.mcna.2020.10.002&rft_id=info:pmid/33589107&rft.issn=0025-7125&rft.volume=105&rft.issue=2&rft.spage=341&rft.pages=341-353&rft.date=2021&rft.jtitle=Medical+Clinics+of+North+America&rft.atitle=Pregnancy+and+Management+in+Women+with+Rheumatoid+Arthritis%2C+Systemic+Lupus+Erythematosus%2C+and+Obstetric+Antiphospholipid+Syndrome&rft.aulast=Castro-Gutierrez

40. Woods L, Perez-Garcia V, Hemberger M. Regulation of placental development and its impact on fetal Growth—New insights from mouse models. Frontiers in Endocrinology. 2018;9. https://www.frontiersin.org/articles/10.3389/fendo.2018.00570.

41. O’Brien K, Wang Y. The placenta: A maternofetal interface. Annu Rev Nutr. 2023;43(1):301–325. 10.1146/annurev-nutr-061121-085246. doi: 10.1146/annurev-nutr-061121-085246.

42. Danish Agency for Labour Market and Recruitment. Sickness benefits for a salary earner. https://lifeindenmark.borger.dk/working/work-rights/leave-of-absence/sickness-benefits/sickness-benefits-for-a-salary-earner Web site. Updated 2024. Accessed Feb 1, 2024.

