## Supplementary material for "Fetal Growth Associated with Maternal Rheumatoid Arthritis and Juvenile Idiopathic Arthritis": Manusript

Table S1. ICD-10 codes to identify women who were diagnosed with RA and JIA prior to, and during their pregnancies.

| **Condition** | **ICD-10 code(s)** |
| --- | --- |
| Rheumatoid Arthritis | M05.X–M06.X |
| Juvenile Idiopathic Arthritis | M08.X |

Table S2. Anatomical Therapeutic Codes of Antirheumatic Therapies (ART) that were retrieved from the Danish National Prescription Registry (NPR). *

| **Medications** | **ATC Code(s)** |
| --- | --- |
| Glucocorticoids | H02AB |
| Prednisone | H02AB07 |
| Betamethasone | H02AB01 |
| Hydrocortisone | H02AB09 |
| Non-steroidal anti-inflammatory drugs | M01A |
| Methotrexate | L01BA01, L04AX03 |
| Hydroxychloroquine | P01BA02 |
| Leflunomide | L04AA13 |
| Sulfasalazine | A07EC01 |

Table S3. Estimated fetal weights and birth weights of offspring among women with and without RA/JIA.

HCQ: hydroxychloroquine; SSZ: sulfasalazine

± Corticosteroids (CS): betamethasone, prednisone, and hydrocortisone.

Reference group*: women without RA/JIA

* Mean difference in Z-scores between women with RA/JIA and women without RA/JIA (reference group).

** Model adjusted for maternal age, maternal BMI, smoking status, income status, race, parity, birth year, pre-pregnancy hypertension, pre-pregnancy diabetes, and co-medication use.

| Estimated Fetal Weight (Z-score) During Second Trimester | | | | | | | | Birth Weight (Z-score) | | | | | | | |
| --- | --- | --- | --- | --- | --- | --- | --- | --- | --- | --- | --- | --- | --- | --- | --- |
|  | Mean value | Crude model | | | Adjusted model** | | |  | Mean value | Crude model | | | Adjusted model** | | |
|  |  | Mean  Difference* | 95% CI | p-value | Mean  Difference | 95% CI | p-value |  |  | Mean  Difference* | 95% CI | p-value | Mean  Difference | 95% CI | p-value |
| No RA/JIA  (Ref) | Ref | - | - | - | - | - | - | No RA/JIA | Ref | - | - | - | - | - | - |
| RA/ JIA | 0.07 (0.02, 0.11) | 0.05 | 0.00,  0.09 | **0.031** | 0.05 | 0.01,  0.10 | **0.022** | RA/ JIA | -0.14  (-0.19,  -0.10) | -0.07 | -0.11,  -0.03 | **0.001** | -0.08 | -0.13,  -0.04 | **<0.001** |
| **Antirheumatic medications use during pregnancy among women with RA/JIA compared with the reference group*** | | | | | | | | | | | | | | | |
| CS^±^ | -0.03  (-0.16, 0.10) | -0.05 | -0.18, 0.08 | 0.5 | -0.05 | -0.18,  0.08 | 0.5 | CS | -0.35  (-0.49, -0.21) | -0.28 | -0.40,  -0.15 | **<0.001** | -0.31 | -0.43,  -0.18 | **<0.001** |
| HCQ | -0.03  (-0.27, 0.20) | -0.05 | -0.27,  0.36, | 0.8 | -0.02 | -0.34,  0.31 | >0.9 | HCQ use | -0.29  (-0.59,  0.01) | -0.22 | -0.53,  0.09 | 0.2 | -0.27 | -0.59,  0.04 | 0.091 |
| SSZ | 0.39  (0.25, 0.53) | 0.37 | 0.24,  0.50 | **<0.001** | 0.38 | 0.24, 0.51 | **<0.001** | SSZ | -0.26  (-0.39, -0.12) | -0.18 | -0.32,  -0.05 | **0.006** | -0.23 | -0.37,  -0.10 | **<0.001** |

Table S4. Effect analysis of the influence of rheumatoid arthritis (RA)/ juvenile idiopathic arthritis (JIA) and pre-eclampsia (PEC), as a mediator on small for gestational age (SGA).

| Mediator | Total Effect Odds Ratio (OR)* | Controlled Direct Effect OR** | Proportion Mediated (%) ^¶^ |
| --- | --- | --- | --- |
| PEC ^±^ | 1.47 (1.13-1.79) | 1.45 (1.22 – 1.83) | 4.23 |

* Effect of RA/JIA on SGA

** Effect of RA/JIA on SGA that is not mediated by PEC

± Effect of RA/JIA on SGA mediated by PEC

¶ Proportion of the effect of RA/JIA on SGA, mediated by PEC: calculated as log(indirect effect)/log(total effect)*100

± Adjusted for age, BMI, smoking status, income status, race, and parity (+ pre-gestational hypertension and pre-gestational diabetes between mediator and outcome)
